# A Mummers Farce - Retractions of Medical Papers conducted in Egyptian Institutions

**DOI:** 10.1101/2023.02.20.23286195

**Authors:** Rahma Menshawey, Esraa Menshawey, Bilal A. Mahamud

## Abstract

**Rationale and Objective:** Egypt currently holds a record for the most retractions in the continent of Africa according to the Retraction Watch database, and the 2^nd^ highest of countries in the Middle East. The purpose of this study was to perform a specific analysis on retracted medical publications from Egyptian affiliations to outline or delineate specific problems and solutions.

**Materials and Methods:** The Retraction Watch Database, Google Scholar, SCOPUS, PubMed, and journals sponsored by the Egyptian Knowledge Bank were searched for all Egypt affiliated retracted medical publications up to the date of August 31^st^ 2022. We observed for the reason(s) for retraction, number of citations, the length of time between publication and retraction and more.

**Results:** 68 retractions were identified that could be linked directly to a known Egyptian institution listed in the study methodology. Most retractions originated from the speciality of Obstetrics and Gynecology (n=22), followed by Anesthesia (n=7). The top 3 reasons for retraction included unreliable results, FFP level misconduct, and duplicate publication. The number of retractions significantly increased over the years, especially in 2022. When taking into account the number of medical publications per institution, the institute with the highest rate of retractions was Mansoura University, while the lowest rate was Cairo University.

**Conclusion:** The number of retracted medical Egyptian publications continues to increase over time, as more issues are uncovered in research coming from this region. Medical papers from this area have been the focus of investigations that have suggested that many results are statistically unlikely to have occurred. Authors must employ a higher ethical standard in their work, while institutions must be openly collaborative with investigations and enact penalties where needed to deter future misconduct. Future studies on retracted articles should employ a methodology that considers the institutions where the studies were conducted in order to obtain a better understanding of specific problems in certain countries or regions.

---

> “Falsehood flies, and truth comes limping after it, so that when men come to be undeceived, it is too late; the jest is over, and the tale hath had its effect: like a man, who hath thought of a good repartee when the discourse is changed, or the company parted; or like a physician, who hath found out an infallible medicine, after the patient is dead.”
>
> – **Jonathan Swift**

## Introduction

Scientific misconduct, once identified, can result in the retraction of published work. Misconduct includes fabrication of data, falsification of data, and plagiarism as defined by the US Office of Research Integrity(Gross, 2016; Resnik, 2019a; Resnik et al., 2015). Misconduct has devastating consequences as the falsified results can have major impact on clinical guidance and decision making, and therefore on the lives and well being of our patients(Steen, 2012). Once a scientific publication is identified to have this level of academic dishonesty, it may be retracted from the literature, reported to the affiliation of the authors, and even result in criminal prosecution if substantial levels of fraud and harm have resulted from the misconduct(Bülow & Helgesson, 2019; Leung, 2019).

We embarked on this study to examine solely the retraction of Egyptian medical papers in order to delineate causes of retraction.

## Methodology

In this cross-sectional study, we examined medical papers found on PubMed, Scopus and Google scholar using the search query “Egypt”, and “Retracted”, or “Retraction Notice”. We also used the Retraction Watch database to identify papers retracted from the country listed “Egypt” on their database (http://retractiondatabase.org/).We also examined the Egyptian Journals collection published by Springer Nature which is sponsored by the Egyptian Knowledge Bank as it is likely Egyptian authors are submitting to these journals, and some of which are not Scopus or PubMed Indexed.

(https://www.springer.com/gp/campaign/egyptian-journals-ekb)

A total of 9954 papers yielded from the search results were examined according to our inclusion criteria. Any duplications were removed.

To be included in our study as an “Egyptian” paper, Egyptian affiliation must be specifically listed in:

- Any mention of an Egyptian affiliation referred to in acquiring ethical approval for the study, or listed as the place where the study took place in the *methods and or acknowledgements* section of the manuscript,

Only Original Research or Case reports or series were included-no reviews, editorials, grey literature or otherwise was considered. Papers with any expression of concern were not included in this study. We included any retracted paper until the date of Aug 31^st^ 2022, that was still available in its full-text form.

Outcomes collected included: the date the paper was published, the date the paper was retracted, the number of authors, subject area, whether or not the paper was COVID-19 related, the number of reads or accesses of the paper, the number of citations, the number of Tweets received by the papers and the upper bound followers (these metrics were either listed directly by the journal, or hosted by PlumX Metrix, or Altmetric), the Journal, Publisher and Cite Score of the retracted paper, the affiliation of the study, reason(s) for retraction, and whether or not the authors approved the decision.

The number of citations were determined using Google Scholar (and or Web of Science).

Causes or reasons for retractions were determined from the retraction notices and broadly categorised as:

- Unreliable results – any mention of data errors, or inconsistencies, methodological errors, or suspicions into the integrity of the presented data
- Duplication – if the work had in full or in part been previously published
- Failure to provide data – failure to provide data sheets upon request for investigation
- Plagiarism – false attribution of another persons work as one’s own
- Fabrication or Falsification – misrepresentation of results (including manipulation – of images or otherwise)
- Consent concern – failure to obtain written informed consent by patient participants.
- Author dispute
- Author request
- Fake authorship
- Fake peer review
- No institutional board approval
- Publication error
- Unspecified causes – no clear mention of the reason for retraction in the retraction notice
- Other causes

Descriptive analysis was reported as averages and standard deviation or as median and range when appropriate.

P values <0.05 were considered significant. Spearman Rho was used to determine if there was significant relation between two variables. Statistical analysis was conducted using both Microsoft Excel and MedCalc for Windows version 19.1 (MedCalc Software, Ostend, Belguim)

## Results

A total of 68 retracted publications met our inclusion criteria. The time from publication to retraction was a median of 698.5 days (IQR, 1322), or 1.91 years. Figure 1 shows the days between publication and retraction of the articles. The shortest and longest time to retraction was 42 days and 5054 days respectively.

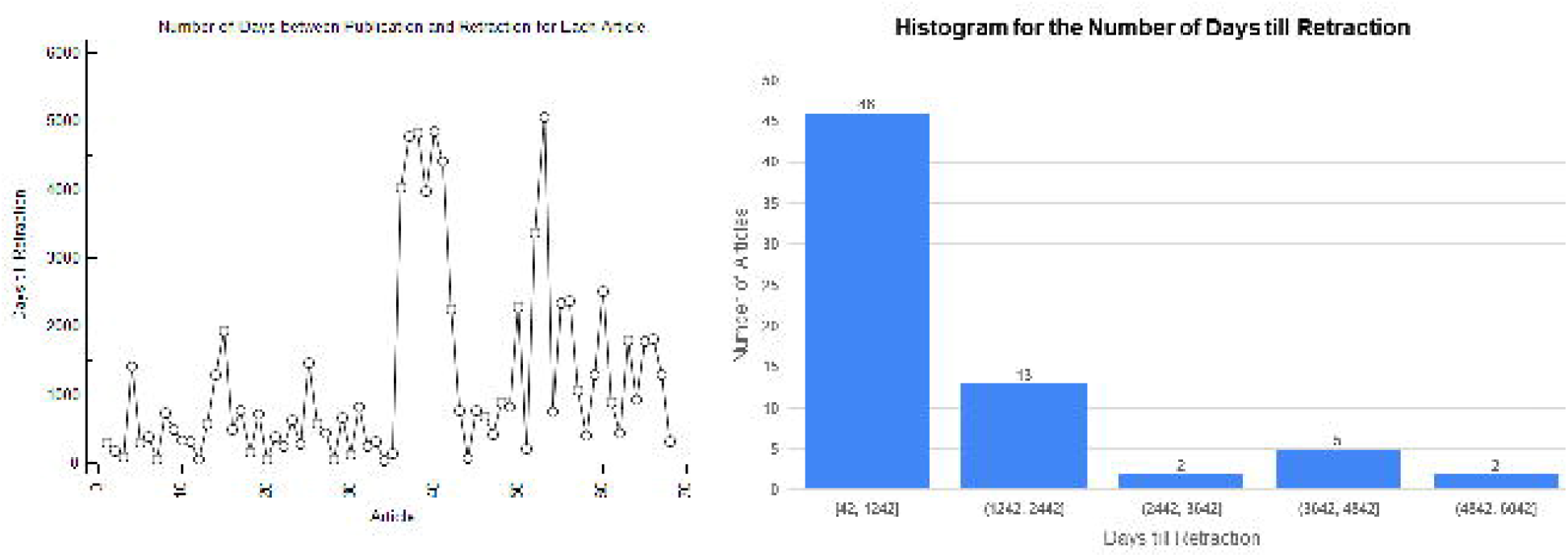

Retractions were highest in 2022 which encompassed 35.24% of the identified retracted papers (n=24). The number of retractions have increased over the years, significantly (Spearman Rho = 0.616, P=0.0191, 95% CI for rho = 0.126 to 0.864) (see figure 2).

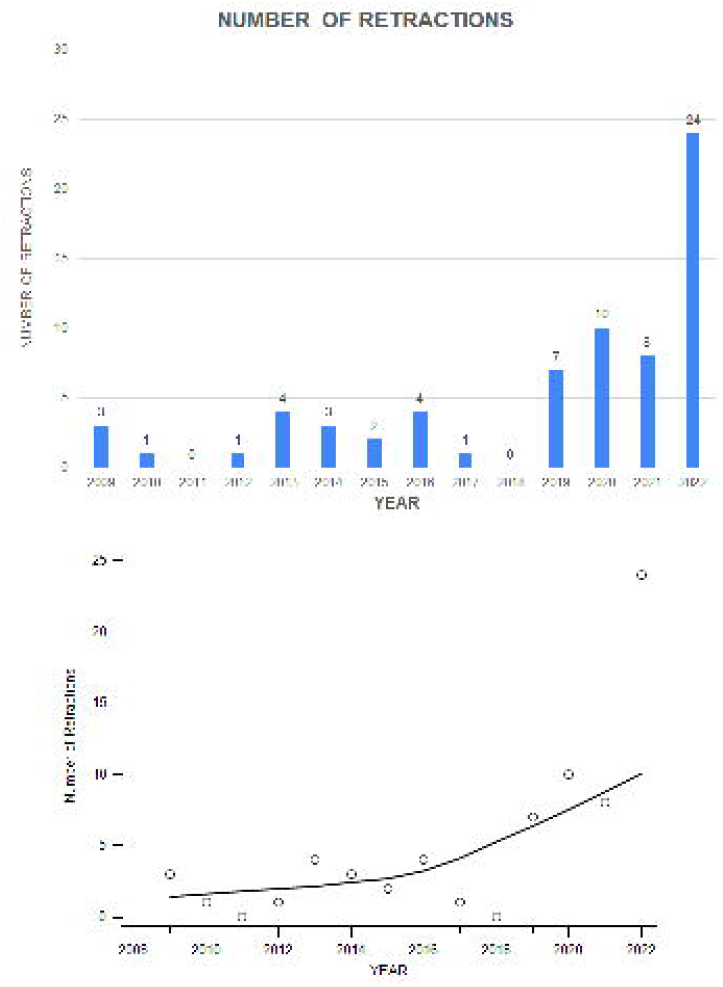

The median number of authors on the retracted papers was 4 (IQR, 2) with the least number of authors being 1 and the highest number of authors was 17.

Retractions were highest in the field of Obstetrics and Gynecology, 32.35% (n=22), and in Anesthesia, 10.29% (n=7) (see figure 3). 7.35% of retracted articles were COVID-19 related, (n = 5).

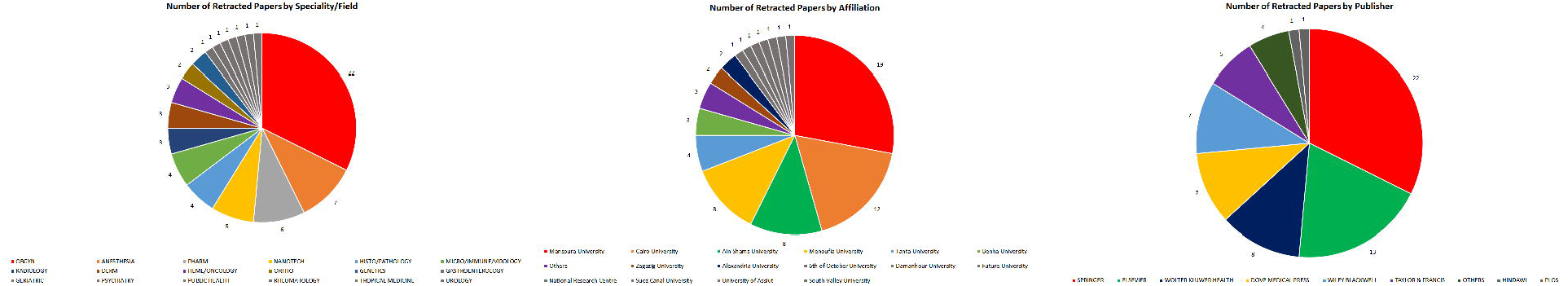

The median number of citations was 11 (IQR = 30), with the lowest value being 0, and the highest number of citations being 255.

Affiliations with the most retractions were Mansoura University, Cairo University, and Menoufia and Ain Shams University, 27.94% (n=19), 17.64% (n=12), 11.76% (n=8), 11.76% (n=8), respectively.

If we adjust for the number of medical papers published from these institutions (see table 1), then the institution that had the highest rate of retractions was Mansoura University with 0.230% retraction rate, while the affiliation with the least rate of retractions was Cairo University at 0.058%.

The publishers with the highest rates of retractions were Springer, 32.35% (n=22), and Elsevier, 19.11% (n=13). Where the Cite Score was available, the median Cite Score was 4.1 (IQR 3.7), with the lowest and highest scores being 0.70 and 70.2 respectively.

Among the 68 retracted papers, 98 reasons for retraction could be identified (1.44 causes per article) (see table 2, figure 4). Unreliable results, FFP type misconduct, and duplication of publication dominated as the main reasons for retraction. Failure to provide data sheets on the request of the Editor of the journals was another common reason for retraction. Among the FFP type of misconduct, 7 retraction notices mentioned a direct concern for manipulation of data or images. Other ethical concerns included failure to obtain patient consent, and Institutional Board Approval.

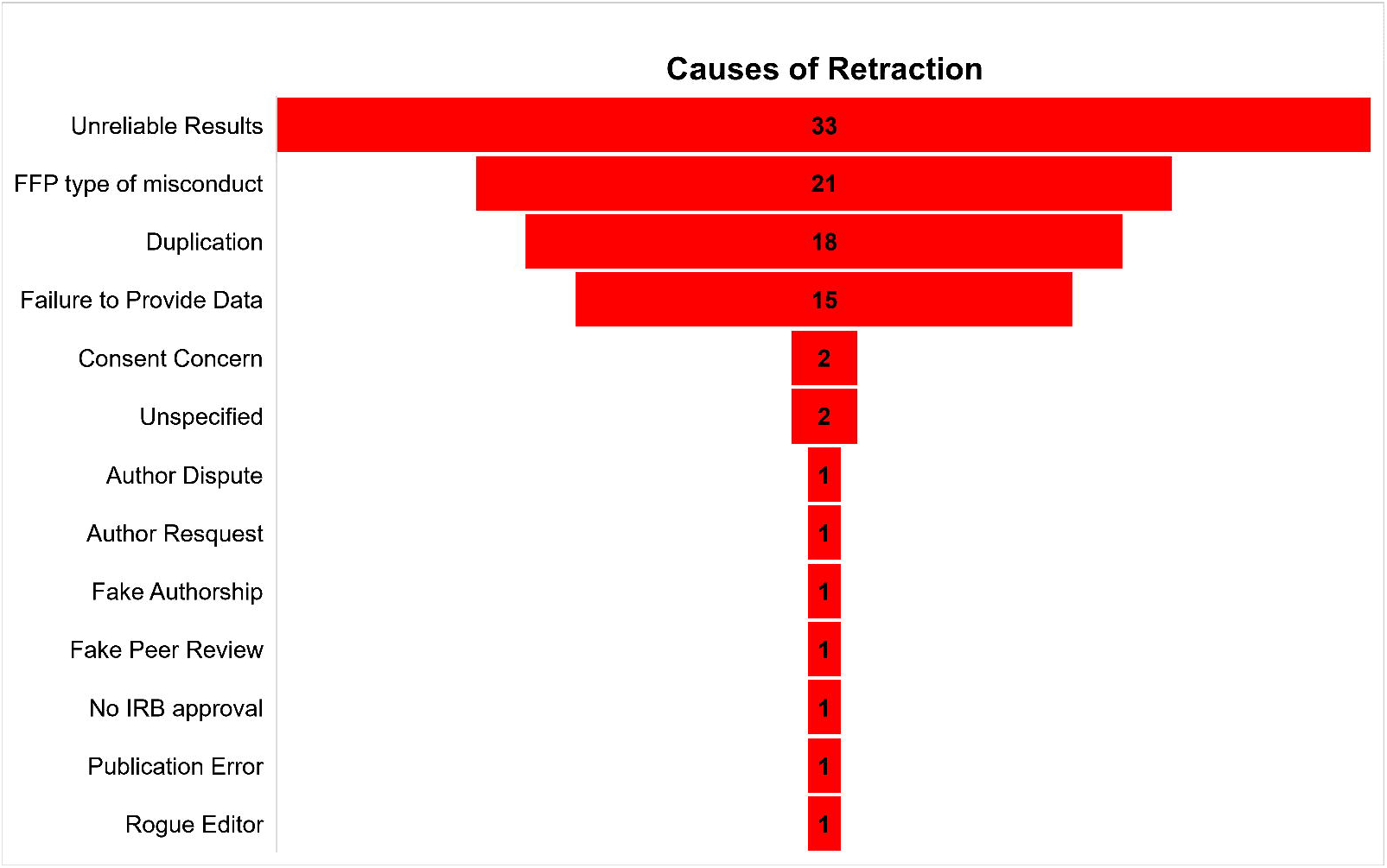

Twenty-seven retraction notices made mention to the authors opinions on the retraction. 9 mentioned disagreement with the decision, 1 mention disagreement while another author did not respond. 12 articles contained total agreement with the decision, while 3 articles showed agreement and other authors not responding. 2 articles showed agreement but the authors disagreed with the wording of the retraction notice.

## Discussion

In the medical field, research and its outcomes can play a pivotal role in the decision-making process of the care of patients. Therefore, it is of utmost importance that research is conducted accurately and to a high degree of ethical standard. Once fabricated, plagiarised, or duplicated work enters the space of the literature, it is likely to skew one decision over another, in an erroneous and potentially detrimental way resulting in harm. Therefore, it is rightful to suggest that the ideal perspective is to recognise that research misconduct is harmful to society as a whole - from the patient level, to the practitioners unknowingly relying on these results, and may dismantle the general trust in healthcare systems. Ultimately, since research is publicly accessible, the potential for harm can spread worldwide (Barde et al., 2020; Gupta, 2013; Rahman & Ankier, 2020).

This exact circumstance was discovered by Bordewijk et al. While incidentally updating a systematic review and meta-analysis they observed unusual similarities in randomised trails conducted by 2 authors from Mansoura University on the topic of ovulation. These similarities could not be accounted for by chance alone and were unlikely to have occurred under any real observable biological reality(Bordewijk et al., 2020). A call to investigate the trials conducted by these authors, as well as any in the region, has led to a domino effect of investigations and retractions of papers from Mansoura University and in the field of Obstetrics and Gynaecology. In our study we recognised these findings as Mansoura University had the highest rate of retracted papers, and OBGYN had the most retractions of any field.

Misconduct in all fields of research can lead to retractions, and this has the power to tarnish the credibility of future work and the reputation of the affiliated institutions. Misconduct maybe commonplace in certain regions or institutions as part of a commonly accepted professional norm, or the belief that there is a low likelihood to be caught or penalised, which calls for strengthened diligence of institutions to foster stronger ethics and enforce administrative punishments when necessary (Okonta & Rossouw, 2014).

Retraction rates are on the rise. Ultimately, there is an ethical duty to report misconduct in all aspects of professional work, and this should be bolstered with a research culture attitude that prioritises ethical conduct and addressing the barriers to reporting concerns when they arise(Satalkar & Shaw, 2018). U.S federal policy defines misconduct as FFP – Fabrication, Falsification and Plagiarism. Some arguments have been made that this definition must be broadened to include failure to disclose conflict of interest, sexual harassment, deceptive use of statistics, and much more(Resnik, 2019b). In our study, we identified several reasons for retractions that extend beyond FFP, including methodological errors, failure to provide complete data sheets to corroborate study results and statistical findings, ethics concerns typically involving failure to obtain consent of study participants, duplicate publication, authorship disputes, and rogue editors and compromised peer review.

Plagiarism is the act of stealing another person’s words or ideas and claiming them as ones own. The research output on identifying plagiarized work in the medical field is steadily rising with alarming findings(Baskaran et al., 2019; Higgins et al., 2016; Menshawey et al., 2022). Plagiarism can be easily avoided by employing original writing skills, complemented with the use of commercially available similarity checking tools to identify problematic areas of the text, by the authors and the journals. Although many journals check submitted work using similarity checking tools, there is room for these results to be misinterpreted if not paired with human judgement (Menshawey et al., 2022; Taylor, 2017).

Duplicate publication was another leading cause of retraction in among the articles we examined. Duplicate publications are another serious and prevalent problem in medical literature (Haworth et al., 2014; Hong et al., 2015; Le et al., 2015). Duplicate publication counts as a redundancy in the scientific literature and are considered highly unethical and are condemned(Haworth et al., 2014). It wastes the time of readers, reviewers, editor and publishers with redundant results, and they further pose issues by skewing the results of metanalysis, and can lead to retractions of those papers that cited them(Bolland et al., 2022; Huston & Moher, 1996; Tramer et al., 1997). Duplicate publications also violate copy-right agreements. One of the retracted papers identified in our study was eventually found to have been previously published in 3 times. In response to this, the journal took swift action:

“The authors were initially served with a show-cause notice and then the dean of the university was also informed when the authors did not respond… a complete restriction on the part of the journal on all future articles in which they are assigned/mentioned as an author/coauthor was imposed and the author was notified accordingly… authors are barred from submitting manuscript(s) to IJD in future” (“Serum Mucosa-Associated Epithelial Chemokine in Atopic Dermatitis: A Specific Marker for Severity: Retraction,” 2014)

Our study aimed at capturing a comprehensive view of retracted papers specifically conducted within Egyptian institutions. Egypt in particular, holds the record for the highest rate of retractions of an African country on the Retraction Watch Database (Nordling, 2019). Therefore, it is imperative that a more specific and selective methodology is utilized to delineate unique issues. There are growing frustrations with the lack of institutional action, response, or even communication with the Editors of journals when concerns arise(Marcus, 2020).

Institutions must be made aware of the impact of retractions on their reputation, as well as develop policies on the matter of misconduct, and strictly enforce swift punitive actions when misconduct is identified-proactive engagement with the Editors of journals and avoidance of obstructing or delaying investigations is necessary (Lievore et al., 2021; Steen et al., 2013; Stern et al., 2014; Yi et al., 2019).

Fake peer review, fake authorship, and even fake editors are a clue to the lengths researchers can go to in order to publish their work. One remarkable example of this, that we examined, involved a rogue editor. This paper, which is related to the COVID-19 topic, was published in an unrelated nanoparticle research journal. The article in question was accepted through a guest-edited special issue. The Editor-in-Chief, observed early on the concerns with the editorial handling and peer review of this special edition and retracted all articles published therein. The article was out of scope with the journal, and the authors disagreed with the decision(Khalifa et al., 2020). Further investigation revealed that the group of academics that had proposed the special issue were being impersonated by an organized group that had purchased email domains similar to those of their affiliated universities, in order to hack the publishing process(Pinna et al., 2020). This is an alarming example of the extent some are willing to go through (developing an entire rogue network) just to get published.

To quote:

“As editors, we are committed since many years to our journal and will continue to invest much of our time to work with authors to promote …the highest standards thanks to the dedicated work of our associate editors and reviewers, but it is also clear that under the present circumstances, it is rarely rewarding and fun.” (Pinna et al., 2020)

Other studies have addressed misconduct in literature in other regions. In Nigeria, one study examined the attitudes and perceptions towards misconduct and found that half of the respondents were aware that a colleague had engaged in misconduct, over 88% were concerned about the level of misconduct in their institution and are worried about its effects and credibility concerns, and the majority believed that getting caught for misconduct was a low chance(Okonta & Rossouw, 2014). If colleagues are aware of misconduct it might be useful for institutions to employ safe unanimous reporting mechanisms for those who wish to disclose their findings without fear of retaliation so investigations can take place when necessary.

Another study examined global scientific retractions from 2001 to 2010 and found higher worldwide average levels of retraction in spite of higher research output in China, India, and South Korea(He, 2013). Trends showed that although scientific research output increased in that decade, the rate of retractions increased by 15.67-fold. The USA had the greatest number of retractions in that decade(He, 2013). A study on the entire Retraction Watch database (2018) showed that in the African region, plagiarism and duplication were the leading causes of retraction, while international collaboration showed fewer retractions for these reasons but had more issues with authorship disputes(Rossouw et al., 2020). A bibliometric analysis in retractions in the Middle East in the last two decades showed that both the number of publications and retractions in this region was significantly increasing. Misconduct was prevalent (79.2%), the majority of which was plagiarism, and most retractions were from the medical field. The countries with the most retractions were Iran, Turkey, and Egypt, with 243, 80, and 58 retractions respectively.

See table 3 (Chauvin et al., 2019) (Cortegiani et al., 2021 (McHugh & Yentis, 2019) (Chambers et al., 2019 (Stavale et al., 2019) (Huh et al., 2016) (Christopher, 2022)) (Rapani et al., 2020) (Gaudino et al., 2021) (Moradi & Janavi, 2018) (King et al., 2018)

Based on our results on Egyptian retractions, focus should be paid to these 3 particular types of misconduct which can be identified – (1) Fabricated/falsified data can be investigated using methods similar to those utilised/devised by Bordewijk et al (Bordewijk et al., 2020), and prompt further investigations when necessary and statistical examinations of data sheets, (2) Plagiarism can be detected by commercially available tools, and pairing text match results with human judgement to avoid misinterpretation(Menshawey et al., 2022), (3) Text matching tools can also be used to locate duplicate publications.

Future research on retracted publications should aim at delineating specific concerns in each region to provide a more specific measure for problematic areas, by taking into consideration the institutions in which the research was conducted.

## Conclusions

Retractions are rising significantly for medical papers conducted in Egyptian institutions. Authors should engage a higher ethical standard when conducting their research, while institutions must develop policies and renewed attitudes against misconduct. More research is needed to understand the root causes or risk factors for misconduct in this region. A revitalised research culture that promotes ethics and no a tolerance attitude towards misconduct is swiftly needed in this region to restore trust on the international scientific stage.

## Data Availability

All data produced in the present study are available upon reasonable request to the authors

## Declarations

### Ethics Approval and Consent to Participate

Ethics approval was not needed for this manuscript

### Consent for Publication

Not Applicable

### Availability of Data and Material

Data available upon reasonable request

Competing interests

The authors declare that they have no competing interests.

### Authors Contributions

All of the authors have contributed to this paper in accordance with ICJME guidelines. RM EM & BA contributed in the data collection, statistics, writing and revising the final draft of this manuscript.

## Acknowledgements

Not applicable

