## Supplementary material for "A Mummers Farce - Retractions of Medical Papers conducted in Egyptian Institutions": table 1

**Table 1: Retraction rates of Egyptian Institutions**

| **Top 5 Affiliations** | **Number of Medical Publications (SCOPUS)*** | **Number of Retractions Identified** | **Rate of Retractions** |
| --- | --- | --- | --- |
| Mansoura University | 8612 | 19 | **0.230%** |
| Cairo University | 20506 | 12 | **0.058%** |
| Ain Shams University | 12186 | 8 | 0.065% |
| Menoufia University | 3598 | 8 | 0.222% |
| Tanta University | 4175 | 4 | 0.096% |
