## Supplementary material for "A Mummers Farce - Retractions of Medical Papers conducted in Egyptian Institutions": table 2

Table 2: Causes of Retraction

| **Causes of Retraction** | **Number of papers retracted** |
| --- | --- |
| Unreliable Results | 33 |
| Duplication | 18 |
| Failure to provide Data | 15 |
| Fabrication/Falsification | 11 |
| Plagiarism | 10 |
| Consent Concern | 2 |
| Unspecified | 2 |
| Author Dispute | 1 |
| Author Request | 1 |
| Fake Authorship | 1 |
| Fake Peer review | 1 |
| No IRB approval | 1 |
| Publication Error | 1 |
| Rogue Editor | 1 |
