## Supplementary material for "A Mummers Farce - Retractions of Medical Papers conducted in Egyptian Institutions": table 3

| Author: | Year: | Field or Country of retracted studies: | Number of Retracted articles: | Main reason for Retraction: |
| --- | --- | --- | --- | --- |
| Chauvin et al.[1] | 2019 | Emergency Medicine | 28 | Plagiarism |
| Cortegiani et al.[2] | 2021 | COVID-19 | 45 | Incorrect results |
| McHugh & Yentis[3] | 2018 | Anaesthesiology | 16 | Data Fabrication |
| Chambers et al.[4] | 2019 | OBGYN | 176 | Plagiarism and Data falsification |
| Stavale et al.[5] | 2019 | Health/ Life Sciences | 65 | Plagiarism and missing data |
| Huh et al.[6] | 2016 | Korean Medical Journals | 111 | Duplicate publication |
| Christopher[7] | 2022 | Veterinary Medicine | 242 | Plagiarism, duplicate publication, Data Fabrication |
| Rapani et al.[8] | 2020 | Dentistry | 180 | Plagiarism, duplicate publication, Data Fabrication |
| Gaudino et al.[9] | 2021 | Biomedical sciences | 5209 | Plagiarism, duplicate publication, Data Fabrication |
| Moradi & Janavi [10] | 2018 | Iranian Scientific Articles | 103 | Plagiarism |
| King et al.[11] | 2017 | Surgical Literature | 184 | Duplication, Plagiarism, Review board Violations |

Table 3: Summary of literature reporting on Retracted papers
